# Association of Animal Species Ownership with under-5 Diarrhea Prevalence in sub-Saharan Africa: An Analysis of 114 Demographic and Health Surveys

**DOI:** 10.64898/2025.12.22.25342862

**Authors:** Steven Sola

## Abstract

Deaths due to diarrhea among children younger than five have been rapidly decreasing, from an estimated 484,781 in 2010 to 245,966 in 2021. A report by the International Livestock Research Institute in 2012 found that the top 13 most important zoonoses were responsible for 2.2 million human deaths. Questions remain whether the benefits of animal ownership outweigh the potential harms of being exposed to potentially deadly zoonotic pathogens. This study uses national surveys to assess the association between animal ownership and under-5 diarrhea in sub-Saharan Africa. Our outcome was a two-week reported prevalence of diarrhea. Information about animal ownership, socio-economic status (SES), age of children, and the family’s primary water source were used in this analysis. We used Poisson regression with a log link to output prevalence ratios from a mixed-effects model.

The study population included 680,752 children under the age of five who lived in rural areas in sub-Saharan Africa between 2005-2022. Our model showed a slight increase of two-week reported diarrhea among children whose families owned animals (PR: 1.02, 95% CI: 1.00, 1.04). Families who owned more animals reported a lower diarrhea prevalence (11.22%) compared to families who owned fewer animals (13.71%). Additionally, children whose families owned no animals had a diarrhea prevalence of 12.09%, compared to children whose families owned more than one species of animal (12.20%). Families who only owned pigs had a diarrhea prevalence of 15.31%.

This study continues to highlight the complicated relationship between animal ownership and reported diarrhea prevalence. Further research needs to examine the role of animal ownership in the context of local weather patterns, especially after extreme precipitation events following extended dry period. There remains an open question whether the benefits of animal ownership outweigh the potential pitfalls, especially in consideration of climate change.

## Introduction

According to the 2021 Global Burden of Diseases Study, diarrhea contributed to an estimated 340,429 deaths among children younger than five years old, which was a decrease from 552,137 deaths in 2015 and 771,312 deaths in 2010 [1]. Specifically among children less than five years old in sub-Saharan Africa, there were an estimated 245,966 deaths in 2021, 365,584 deaths in 2015, and 484,781 deaths in 2010, with unequal geographic distributions of diarrheal deaths throughout sub-Saharan Africa[1]. Despite this trend of decreasing deaths due to diarrheal diseases, sub-Saharan Africa continues to experience the highest rates of diarrheal disease globally [1], and those living in lower socioeconomic conditions are disproportionately affected by these diseases [2,3]. Further efforts are needed to reduce the burden of diarrheal diseases among these populations, especially in sub-Saharan Africa.

Zoonotic infections, which are infections that are passed from animals to humans, can be transmitted via direct contact, soil, air, or water. According to the World Health Organization (WHO), approximately 60% of the 1400 infectious disease pathogens are zoonotic [4]. A recent report from WHO Africa identified a 63% increase in zoonotic outbreaks from 2012-2022 compared to 2001-2011 [5]. This same report hypothesized the increase is due to an increase in food derived from animals, increased urbanization, and increased mobility across the continent [5], although this increase could also be the result of an increase in molecular diagnostics worldwide. A report by the International Livestock Research Institute in 2012 found that the top 13 most important zoonoses, such as generalized gastrointestinal pathogens, Leptospirosis, Hepatitis E, and Tuberculosis, were responsible for 2.2 million human deaths in 2.4 billion cases of illness, with generalized zoonotic diarrhea having the highest burden of disease [6].

Given the potential risk from zoonotic disease exposure, understanding the role of peridomestic animals in diarrheal outcomes among children under five years old is important. Animal husbandry provides benefits to families throughout sub-Saharan Africa by giving families an additional source of income [7], empowering women [8,9], and increasing nutrition for the family [10]. A systematic review by Zerfu et al. shows that livestock keeping can increase height-for-age and weight-for-length Z scores, but it can also be associated with increased morbidity and mortality [11]. A prior study using data from the DHS across sub-Saharan Africa identified a null association between animal ownership and the two-week prevalence of diarrhea [12].

However, there was significant heterogeneity of findings among the countries included in the surveys at the national level: Ten of the countries showed protective effects of animal ownership and diarrhea prevalence, six showed no effect, and 14 showed harmful associations [12].

The goal of this research was to extend the analysis of this previous study and examine the associations between 2-week prevalence of diarrhea and other demographics, such as the total number of animals owned, socioeconomic status (SES), and the types of animals to which a child may be exposed, as the previous study only considered a family’s ownership of poultry and not other animals. A secondary goal was to evaluate the ownership of multiple species of animals with the 2-week prevalence of diarrhea, and compare it with households that only owned a single species of animal. The debate about the cost-benefit ratio of animal husbandry, especially in rural areas, is ongoing. It is unknown whether certain types of animals, such as poultry, ruminants, or equids, would be more beneficial or harmful to people in these areas. We address this gap in this research by using a large dataset made available by the DHS over 18 years and 114 individual surveys.

## Methods

### Study Design

To estimate the association between animal ownership and under-5 diarrhea, we leveraged the United States Agency for International Development’s (USAID) DHS data, similar to prior work. DHS household surveys provide nationally representative information about household composition, health, and other factors [13].

### Survey Data

For this analysis, our exclusion criteria were: all individuals who lived in urban or unknown urbanicity settings, according to DHS definitions [14]. We also excluded everyone five years of age and older, those who had a missing age, or children who were already deceased.

Additionally, we excluded all data that was recorded before 2005 because the DHS did not start collecting information about animal ownership until that time. The result was a dataset which we used to examine the association of diarrhea among children aged four and under, stratified by families who did and did not own animals between the years 2005-2022.

### Statistical Analysis

We utilized a mixed-effects Poisson regression model with a log link to assess the association between household animal ownership and the two-week prevalence of diarrhea among children under the age of 5. The results of these models are prevalence ratios. We utilized the *lme4* package in R [15]. Our model was specified as follows:

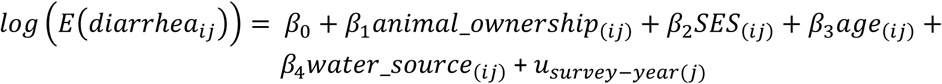

Where:

β_1_ is whether the family owns any animals (dichotomous).

β_2_ is to the SES of the household that the individual lives in (ordinal).

β_3_ is to the age of the child under the age of five (continuous).

β_4_ is whether the family’s primary water source is improved or unimproved (dichotomous).

𝑢 is the random intercept for the survey-year (e.g. “South Africa 2016” or “Senegal 2007”). subscript *i* is used to denote measurements that change on the individual level.

subscript *j* is used to denote measurements that change on the group (i.e. “survey-year”) level.

The primary outcome for our analysis was the variable “H11”, which questions whether the child has had diarrhea within the past two weeks or 24 hours. There were no reports of children experiencing diarrhea in the past 24 hours, so this outcome solely represents the 2-week diarrhea prevalence.

The primary exposure for our analysis asks whether the family, “Owns livestock, herds or farm animals”. While most surveys had a common coding scheme for specific animals, there were some differences between older and newer surveys. As a result, we assessed animal ownership for each individual survey based on that particular survey’s codebook. After extracting those data, we classified each entry into the following categories: chickens, goats, bulls, cattle, horses, pigs, ducks, camels, rabbits, bees, rodents, sheep, and other. We further categorized these into the following categories, which we used for our analyses: poultry (chicken/duck) (avian), goat/sheep (small ruminant), cattle/cow/bull (bovine), horse/donkey (equid), pigs, and other.

The maximum number that a family could report in each of the initial categories was capped at 95 individual animals, per DHS survey protocols. SES (HV270) was recorded for each household on an ordinal scale, where “1” represented the poorest households and “5” represented the richest households. SES was only considered within each survey-year, and not across all the surveys included in the analysis. Information about the primary source of water for each household (HV201) was extracted from each survey. We used three categories for this study: unimproved (“surface water” and “unimproved” JMP categories), improved (“limited”, “basic”, and “safely managed” JMP categories), and unknown/other [16].

We included the fixed-effects for SES (ordinal), age of child (continuous), and water source (dichotomous) based on previous research that has included these covariates in their models [2,3,12,17–19] and their potential to be confounding in the relationship between animal ownership and 2-week prevalence of diarrhea. We considered inclusion of fixed effects for head-of-household education and the gender of the child, and random intercepts for household and cluster, but ultimately excluded these as they did not significantly change the results from the model and to keep the model parsimonious.

We also assessed whether the number of animals in the household was associated with the diarrhea prevalence of children. We split the number of children aged four and under with diarrhea into quintiles and then assessed the average number of animals within each quintile, along with the associated diarrhea prevalence. A Pearson’s correlation test was used to assess a correlation between the number of animals that a family owns and the average SES within that group. A chi-square test of independence was used to assess a statistically significant difference in the different categories of animal ownership among children who were and were not reported to have a 2-week prevalence of diarrhea. A Kruskal-Wallis rank sum test was used to assess a statistically significant difference in the different categories of animal ownership among children and their family’s reported SES. Finally, a Cochran-Armitage test for trend was used to assess a statistically significant trend in the different categories of SES among children who were and were not reported to have a 2-week prevalence of diarrhea.

All analyses were completed using R 4.2.1 [20]. Replication code is made publicly available on https://github.com/sqsola/ClimateWASH. Data can be obtained through a request to the Demographic and Health Survey team at https://dhsprogram.com/. Permission to download data was received prior to commencing data analyses.

## Results

### Datasets

Our source dataset included 12,007,303 individuals across 172 survey-years. From our source dataset, we excluded all individuals who lived in urban (n = 3,116,550) or had an unknown urbanicity (n = 1,747,960). We further excluded everyone five years of age and older (n = 1,614,644), those who had a missing age (n = 4,742,118), or children who were already deceased (n = 259). Finally, we excluded all children before 2005 due to no information regarding animal ownership being available (n = 105,114). A total of 680,752 children aged four and under were included in our analysis (see figure 1). Our dataset encompassed 114 survey-years from 35 countries (see supplemental table 1).

**Figure 1.**
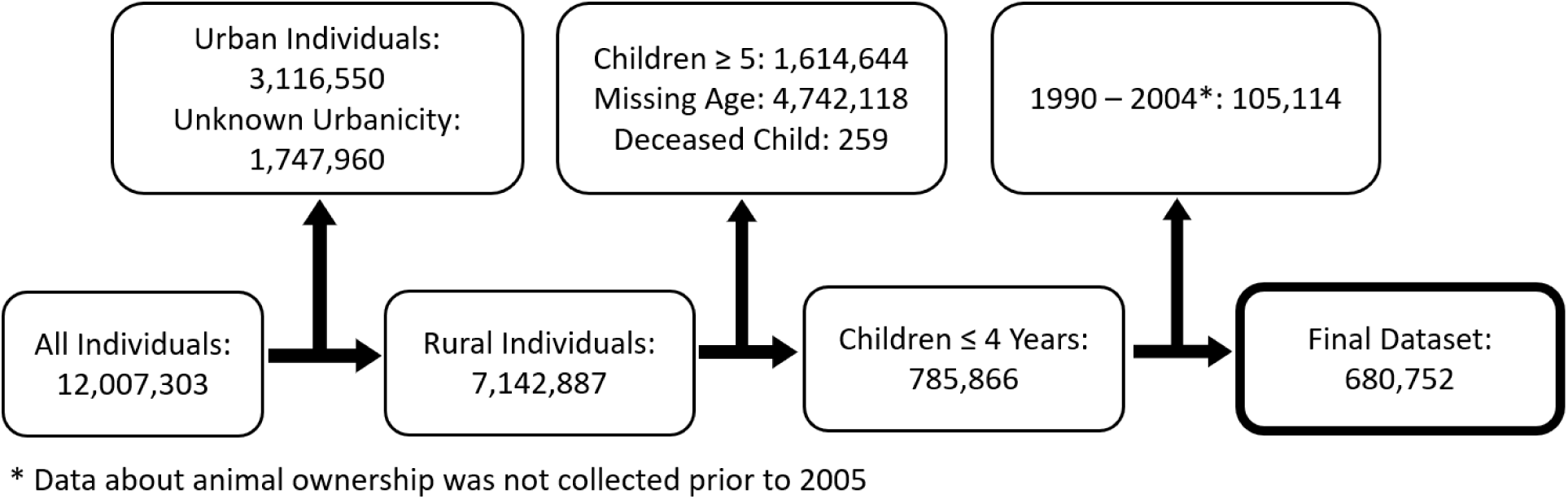
Inclusion of individuals into the dataset used for this analysis. The final dataset included 680,752 children under the age of five between the years 20005-2022.

### Analyses

There were 423,224 children aged four years old and younger whose families reported animal ownership (62.2% of all children). The overall prevalence of diarrhea in our study was 11.84% (see table 1). Among these children whose families own animals, 52,389 children were reported to experience diarrhea within the previous two weeks (12.38% prevalence). There were 173,587 children whose families did not own animals, of which 20,981 children in this group were reported to have diarrhea in the previous two weeks (12.09% prevalence). There were 83,941 children who lived in households with an unknown number of animals, of which 7,228 of these children were reported to have diarrhea (11.61% prevalence).

**Table 1.** Diarrhea prevalence stratified by the number of animals that a household owns. Categories were created by taking the number of children under the age of five with diarrhea and creating five categories.

| **Number of Animals in Household** | **Children Under 5** | **Children Under 5 with Diarrhea** | **Average Number of Animals per Child under 5** | **Average SES per Child under 5<sup>1</sup>** | **Diarrhea Prevalence (%)** |
| --- | --- | --- | --- | --- | --- |
| Unknown | 83,941 | 7,228 | Unknown | 2.37 | 8.61 |
| No Animals | 173,587 | 20,986 | 0 | 2.35 | 12.09 |
| 1-3 | 77,639 | 10,648 | 2.02 | 2.26 | 13.71 |
| 4-7 | 87,374 | 11,161 | 5.38 | 2.28 | 12.77 |
| 8-13 | 85,431 | 10,514 | 10.25 | 2.26 | 12.31 |
| 14-25 | 84,496 | 10,157 | 18.65 | 2.22 | 12.02 |
| 26+* | 88,284 | 9,909 | 54.83 | 2.25 | 11.22 |
| Total | 680,752 | 80,603 | 11.63<sup>#</sup> | - | 11.84 |

<sup>1</sup> SES is an ordinal variable with the following levels: 1 = Poorest, 2 = Poor, 3 = Middle Class, 4 = Rich, 5 = Richest
\*The maximum amount of animals within a single category was 95. As a result, it is possible that some households owned more animals than what was reported in the data.
<sup>#</sup> Total number of animals in this study (7,918,898) divided by the number of children under the age of 5 (680,752). As a result, this number represents the average number of animals that each child was exposed to.

Among families who reported animal ownership, children in households who owned 1-3 animals had the highest diarrhea prevalence (13.71%), while children in households who owned 26+ animals had the lowest diarrhea prevalence (11.22%). Children in households who had an unknown number of animals had a diarrhea prevalence of 11.61% (see Table 1). The result of the Pearson’s correlation test between the number of animals that a family owns and the average SES within that group was statistically significant (Pearson’s r = 0.37, p-value = <0.001) (see supplemental figure 1).

In addition to the total number of animals that a household owns, we examined the specific species of animals that the household owns. The highest diarrhea prevalence is in households who only own pigs (15.31%), followed by goats or sheep (13.85%), but these were less commonly kept species. Households who reported owning more than one species of animal had a modest diarrhea prevalence of 12.20%—higher than households without animals but lower than other categories of single-animal ownership. Households who reported owning other animals (such as rabbits, rodents, camels, and bees) had a diarrhea prevalence of 7.16%, although these animals are not known to be major sources of zoonotic diarrheal pathogens (see table 2. The result of the chi-square test of independence to assess a statistically significant difference in the different categories of animal ownership among children who were and were not reported to have a 2-week prevalence of diarrhea test was statistically significant (χ^2^ = 197.9, p-value: <0.001). The result of the Kruskal-Wallis rank sum test used to assess a statistically significant difference in the different categories of animal ownership among children and their family’s reported SES was statistically significant (χ^2^ = 749.8, p-value: <0.001). The different combinations of household ownership of animals is visualized in supplemental figure 2.

**Table 2.** Diarrhea prevalence stratified by the type of animals that a household owns.

| **Single Animal Category** | **Children Under 5** | **Children Under 5 with Diarrhea** | **SES<sup>1</sup>** | **Diarrhea Prevalence (%)** |
| --- | --- | --- | --- | --- |
| >1 Species | 281,369 | 34,379 | 2.24 | 12.20 |
| No Animals | 173,587 | 20,986 | 2.35 | 12.09 |
| Poultry (Chicken/Duck) Only | 89,747 | 11,160 | 2.29 | 12.43 |
| Unknown | 83,941 | 7,228 | 2.37 | 11.61 |
| Goat/Sheep Only | 24,751 | 3,429 | 2.22 | 13.85 |
| Bull/Cow/Cattle Only | 15,134 | 1,950 | 2.42 | 12.88 |
| Horse/Donkey Only | 4,368 | 596 | 2.06 | 13.64 |
| Other* | 4,021 | 288 | 2.29 | 7.16 |
| Pig Only | 3,834 | 587 | 2.48 | 15.31 |
| Total | 680,752 | 80,603 | - | 11.84 |

<sup>1</sup> SES is an ordinal variable with the following levels: 1 = Poorest, 2 = Poor, 3 = Middle Class, 4 = Rich, 5 = Richest
\* Includes rabbits, rodents, camels, and bees

**Table 3.** Diarrhea prevalence stratified by SES status.

| **SES<sup>1</sup>** | **Children Under 5** | **Children Under 5 with Diarrhea** | **Diarrhea Prevalence (%)** |
| --- | --- | --- | --- |
| Poorest | 224,881 | 28,804 | 12.81 |
| Poor | 186,001 | 22,321 | 12.00 |
| Middle | 146,399 | 16,522 | 11.29 |
| Rich | 93,260 | 10,018 | 10.74 |
| Richest | 30,207 | 2,937 | 9.72 |
| Total | 680,748* | 80,602<sup>#</sup> | 11.84 |

| SES<sup>1</sup> | Children Under 5 | Children Under 5 with Diarrhea | Diarrhea Prevalence (%) |
| --- | --- | --- | --- |

<sup>1</sup> SES is an ordinal variable with the following levels: 1 = Poorest, 2 = Poor, 3 = Middle Class, 4 = Rich, 5 = Richest
\* Excludes 4 children with an unknown SES

Lastly, we assessed the diarrhea prevalence stratified by SES category. The highest diarrhea prevalence was among the households with the lowest SES (12.81%), while the lowest diarrhea prevalence was among the households with the highest SES (9.72%) (see table 5). The result of a Cochran-Armitage test for trend test was statistically significant (z = 20.1, p-value: <0.001).

After conducting our mixed-effects Poisson regression, we found that the prevalence of 2-week reported diarrhea among children whose families own animals was 2% higher compared to children whose families did not own animals (PR: 1.02, 95% CI: 1.00, 1.04), controlling for the age of the child, SES group, and family’s water source. The prevalence of 2-week reported diarrhea decreased by 22% for each additional year of the child’s age (PR: 0.78, 95% CI: 0.77- 0.78) between the years 0 and 4. There was a statistically significant decrease in the prevalence of 2-week reported diarrhea among richer SES groups compared to the poorest SES group. Finally, children whose families used an unimproved water source had a 3% higher prevalence of 2-week reported diarrhea compared to families who used an improved water source (PR: 1.03, 95% CI: 1.01, 1.05).

When animal ownership was stratified by the unique groups of animals (poultry [chicken/duck], goat/sheep, cattle/cow/bull, horse/donkey, and pigs), the covariates of age, SES group, and water source are similar to the model presented in Table 4. The only animal that had a statistically significant difference in prevalence is children whose families own horses compared to those whose families do not own horses (PR: 0.94, 95% CI: 0.92, 0.97), showing a protective effect for horse ownership (see supplemental table 3). Additionally, we created three models for the number of animals that a child’s family owns (per 5 animals, per 10 animals, and per 15 animals) and their association with the 2-week prevalence of diarrhea with the aforementioned covariates. All three of these models showed a very slight but statistically significant decrease in the 2-week reported prevalence of diarrhea (see supplemental table 4).

**Table 4.** Results of the mixed-effects Poisson regression model with a log link.

| Covariates | PR | 95% CI | p-value |
| --- | --- | --- | --- |
| Animal Ownership | 1.02 | 1.00, 1.04 | **0.04** |
| Age of Child (Years 0-4) | 0.78 | 0.77, 0.78 | **<0.001** |
| SES Group |  |  |  |
| Poorest | Ref | Ref |  |
| Poor | 0.94 | 0.92, 0.96 | **<0.001** |
| Middle | 0.88 | 0.86, 0.90 | **<0.001** |
| Rich | 0.84 | 0.82, 0.86 | **<0.001** |
| Richest | 0.74 | 0.71, 0.77 | **<0.001** |
| Water Source |  |  |  |
| Improved | Ref | Ref |  |
| Unimproved | 1.03 | 1.01, 1.05 | **< 0.001** |
| Other/Unknown | 1.00 | 0.89, 1.13 | **> 0.9** |

Abbreviations: PR = Prevalence Ratio, CI = Confidence Interval

## Discussion

This research further reinforces the complicated associations between animal ownership and reported diarrhea prevalence. The results of our mixed-effects Poisson regression model show that animal ownership may be associated with the two-week reported diarrhea prevalence among children under the age of five, but this effect is small at only 2%. This result is similar to the pooled OR result (1.00, 95% CI: 0.99, 1.01) in Kaur et al. [12]. A metanalysis by Zambrano et al. found a much stronger association between animal husbandry and diarrheal illness (pooled OR: 2.73, 95% CI: 1.90, 3.93) [21], while a systematic analysis by Penakalapati et al. suggested a more null association between animal ownership and prevalence of diarrhea [22]. The study by Kaur et al. used a similar dataset as this study, while the studies by Zambrano et al. and Penakalapati et al. used different datasets, which could explain the heterogeneity in the results across the different studies.

Our model additionally showed that children in richer SES groups had statistically significant lower prevalence of an episode of diarrhea in the prior two weeks compared to children in the poorest SES group. This finding is consistent with our unadjusted analysis that showed children in the poorest SES group had a 2-week reported diarrhea prevalence of 12.81 compared to children in the richest SES group have a 2-week reported diarrhea prevalence of 9.72%. These findings are concordant with other studies, as noted in a metanalysis conducted by Azanaw et al. [23], who reported that low SES was a recurring theme of being associated with increased diarrhea prevalence among the research included in the study. An increased SES not only affords families better healthcare and improved sanitation facilities, but it also affects which types of animals that they own. As noted in a research paper by Pica-Ciamarra et al. [24], there is some evidence of a “livestock ladder,” in which the poorest families are mainly focused on keeping poultry, while slightly less poor families are focused on keeping small ruminants and/or pigs, and the more relatively afluent families can afford to keep larger ruminants, such as cattle and buffalo. In this study, there is evidence that the more animals that a family owns, the higher their SES level (see supplemental figure 1). Additionally, there is evidence that families with different SES levels own different types of animals (see table 2). In DHS surveys, animal ownership is not taken into account when constructing the SES variable [25], and the variance inflation factors (VIFs) for our animal ownership models (see supplemental table 4) were close to 1, indicating no multicollinearity between SES and animal ownership in these models.

Our unadjusted analyses identified that children whose families owned animals reported a higher diarrhea prevalence (12.38%) compared to children whose families did not own animals (12.09%). Within these families who own animals, the highest prevalence of diarrhea were among families with fewer animals compared to families with more animals, and children whose families owned between 1-3 animals having a 2-week reported diarrhea prevalence of 13.71% compared to children whose families owned 26 or more animals having a 2-week reported diarrhea prevalence of only 11.22%. Additionally, households who reported owning more than one species of animal reported the lowest prevalence of diarrhea (12.20%), excluding households who only owned “other” animals, such as rodents, rabbits, camels, and bees. In our study, the top three animal ownership combinations among children were those who only owned poultry (n=89,747 [22.6%]), those who owned poultry and goats or sheep (n=56,893 [14.4%]), and those who owned poultry, goats or sheep, and bulls or cows or cattle (n=46,368 [11.7%]) (see supplemental figure 2). These findings show that owning more, and a greater variety of animals, is associated with a decreased prevalence of diarrhea. However, caution must be exercised, as a greater number and variety of animals may also suggest a family with a greater SES.

These findings build upon, and further clarify, previous findings related to the association between animal ownership and diarrhea prevalence. In previous studies, livestock and poultry ownership has been shown to increase weight-for-age Z score measurements in some communities, but it also has been shown to be associated with decreased weight-for-age Z scores in other communities, suggesting that animal ownership has a complicated risk and benefit relationship that depends on community characteristics [26]. A metanalysis from 2014 found that, out of 29 studies included, 20 reported an increase in incidence of diarrhea when exposed to animals [21]. Additionally, a study from Western Kenya found an increased prevalence of *Cryptosporidium* infection and subsequent diarrhea among families who owned cows and other livestock, compared to families who didn’t own livestock [27]. An alternative perspective comes from Krumkamp et al., who found an increased risk of *Cryptosporidium* infection among children under five if they had family members or neighboring children who were also infected, while animals only had a marginal effect on *Cryptosporidium* infection [28]. This led the authors to speculate that *Cryptosporidium* transmission in sub-Saharan Africa may be more anthroponotic, rather than zoonotic [28]. These differing results may be a result of small sample sizes in each study. Animal husbandry practices may vary widely, and only including a small sample of people in each study may bias the results of the studies.

One strength of our study is the large number of households included in the analysis. We were able to assess the diarrheal outcomes of 680,752 children under the age of five across 114 surveys and 35 countries using nationally representative surveys. These surveys have been conducted since 1984 and have a standardized set of protocols. Host countries were allowed to add questions that it finds relevant to their priorities. These data afforded us an opportunity to assess multiple dimensions of the households and children who were included in this study, such as their animal ownership patterns, children’s ages, primary water source, and SES. Another strength of this study is our ability to look at the different species that a household owns and their associated 2-week prevalence of diarrhea. A previous study researched the association between ownership of livestock (cows and goats) and poultry (chickens and ducks) and prevalence of Hepatitis E, but found no statistically significant association [29]. A cross- sectional study from Uganda showed that the more animals that a family owned decreased the prevalence of diarrhea in their children under the age of 5 [30]. Previous studies have looked at the association between density of animals and nearby reported illnesses [31,32], with mixed results. This study improves on these previous ones by not only examining the households that have each species of animals, but also households that own more than one species of these animals, as well as expanding the types of animals (e.g. equids and pigs).

There were also some limitations to our study. First, this study uses cross-sectional data, so causation cannot be ascertained using these data. With these analyses, we are only suggesting associations between aspects of the data. Additionally, a household could only state up to a maximum of 95 of a type of animal. This most severely limits our information regarding the true ownership patterns of chickens, ducks, and other poultry among households, as these are the cheapest animals to own compared to the other four categories of animals we examined in terms of both cost and care [24]. Another limitation was the lack of information about where these animals were housed. Chickens may be raised closer to the home compared to other larger livestock, which may expose young children to more chicken fecal pathogens compared to other animals’ fecal pathogens [33,34]. This works in tandem with the number of a specific animal as well, as a household who owns fewer chickens are more likely to keep them closer to the household as opposed to a household who owns more chickens, or owns a chicken farm [35]. A smaller study from rural central Tanzania suggests that sharing water sources with animals and keeping chickens within the home is associated with increased prevalence of diarrhea [36]. More information about animal husbandry practices is needed to more fully understand the complete relationship between animal ownership and prevalence of diarrhea.

Another limitation for this study is the reporting of diarrhea by the primary caregiver, or knowing the source for the diarrheal infection. It has been well-documented that the reporting of diarrhea by the primary caregiver can be flawed [37–39], particularly in DHS surveys [40]. A study based in Maputo detected more than one enteric pathogen in 86% of children, although diarrheal symptoms were only reported for 16% of those children [41], suggesting that pathogen infection may not be a major role in the reporting of diarrhea on household surveys, or there may be an unintended underreporting of diarrhea among a survey’s participants.

Additionally, exposure to animal pathogens is just a single pathway that may lead to an incidence of diarrhea. Rotavirus and *E. coli* are suspected to be the biggest contributors to diarrheal disease in sub-Saharan Africa [42]. *E. coli* is a well-known zoonotic pathogen, while rotavirus may have less, but still some, zoonotic potential [43,44]. A study by Thystrup et al. in 2024 estimated diarrheagenic *E. coli* as having the highest morbidity and mortality across Africa (see supplemental table 2) [42]. Another metanalysis found that among eight studies, exposure to fecal pathogens of animal origin may or may not impair child growth [22], bringing into question whether a focus on fecal pathogens in the environment is a necessary focus.

Future researchers who examine the associations between animal ownership and under-5 diarrhea have a number of considerations. Rainfall patterns and extreme precipitation events may mobilize fecal pathogens in the environment—or conversely may wash them away [45]— which could modify the associations identified in this work. Another consideration for future research is the composition of the family unit, and assessing who is most responsible for caring for the family’s household animals. It has been documented that even though animal care is typically the responsibility of women and children, they hold a relatively small amount of power when it comes to ownership rights, decision making, and handling the income from the care of these animals [45]. An increase in animal ownership could similarly increase the burden of work for women and children in the household, despite the potential increases in nutrition and status for the family.

Although the question of whether a household’s ownership of animals is beneficial or harmful still needs further research and refinement, this work supports that animal ownership— particularly owning a small number of animals or owning pigs or large ruminants—was associated with diarrhea in children four years of age and younger. A family’s SES was an important factor in this relationship, as higher SES households had children with a lower 2-week prevalence of diarrhea compared to lower SES households. Given emerging pressures on animal ownership under climate change scenarios, including risks posed by precipitation events and heat, small household livestock ownership and husbandry practices will continue to evolve.

Recognition of threats of exposure to zoonotic enteric pathogens needs to be balanced by economic and nutrition supports from animal raising efforts and better understanding of the risks and benefits may help to target interventions.

## Data Availability

All data produced in the present study are available upon reasonable request to the authors

## Acknowledgements

This work was carried out at the Advanced Research Computing at Hopkins (ARCH) core facility (rockfish.jhu.edu), which is supported by the National Science Foundation (NSF) grant number OAC1920103.

---

**Supplemental Figure 1.**
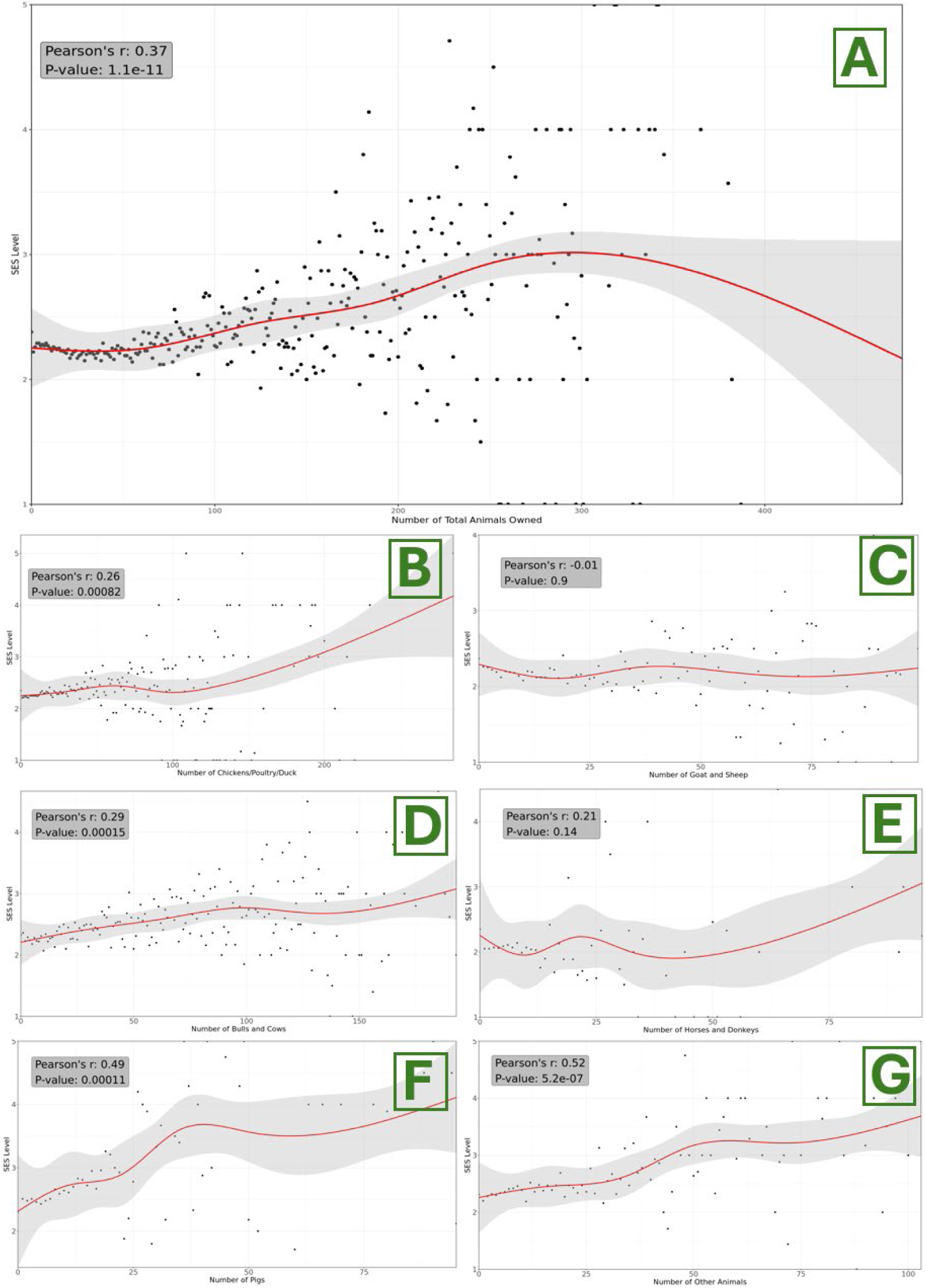
Individuals and their average SES based on animal ownership. All graphs were fit with a natural spline with 5 degrees of freedom. Part (A) shows the overall trend in the number of animals owned and the average SES level of the people who owned that number of animals. For example, all households that owned 1 animal had an average SES level of 2.2. Part (B) shows the number of poultry owned and the average SES level of those individuals. Part (C) shows the number of goat and sheep owned. Part (D) shows the number of bulls and cows owned. Part (E) shows the number of horses and donkeys owned. Part (F) shows the number of pigs owned. Part (G) shows the number of other animals owned.

**Supplemental Figure 2.**
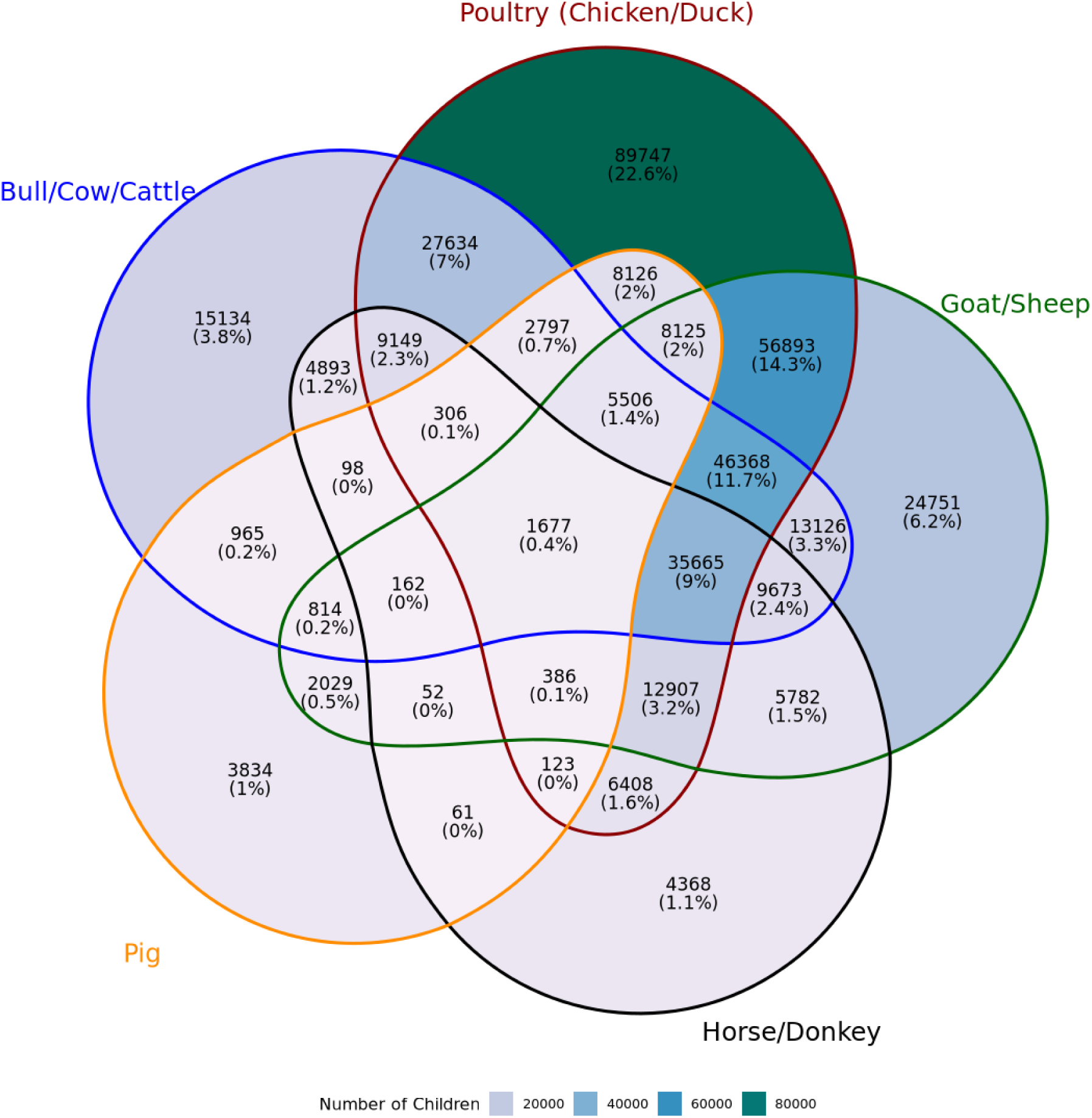
Households and the different combinations of animals owned by each household.

**Supplement Table 1.** Countries and the respective years of their surveys that were used in this study.

| Country | Dataset Years (n = 114) |
| --- | --- |
| Angola | 2006-2007, 2011, 2015-2016 |
| Benin | 2011-2012, 2017-2018 |
| Burkina Faso | 2010, 2014, 2017-2018, 2021 |
| Burundi | 2010, 2012, 2016-2017 |
| Cameroon | 2011, 2018 |
| Chad | 2014-2015 |
| Comoros | 2012 |
| Democratic Republic of the Congo | 2007, 2013-2014 |
| Cote d'Ivoire | 2011-2012, 2021 |
| Eswatini | 2006-2007 |
| Ethiopia | 2005, 2011, 2016, 2019 |
| Gabon | 2012, 2019-2021 |
| Gambia | 2019-2020 |
| Ghana | 2008, 2014, 2016, 2019 |
| Guinea | 2005, 2012, 2018, 2021 |
| Kenya | 2008-2009, 2014, 2015, 2020, 2022 |
| Lesotho | 2009, 2014 |
| Liberia | 2007, 2009, 2011, 2013, 2016, 2019-2020 |
| Madagascar | 2008-2009, 2011, 2013, 2016, 2021 |
| Malawi | 2010, 2012, 2014, 2015-2016, 2017 |
| Mali | 2006, 2012-2013, 2015, 2018, 2021 |
| Mauritania | 2019-2021 |
| Mozambique | 2011, 2018 |
| Namibia | 2006-2007, 2013 |
| Niger | 2012, 2021 |
| Nigeria | 2008, 2010, 2013, 2015, 2018, 2021 |
| Rwanda | 2005, 2007-2008, 2010, 2014-2015, 2019-2020 |
| Senegal | 2008-2009, 2010-2011, 2012-2014, 2015-2016, 2017, 2018, 2019, 2020-2021 |
| Sierra Leone | 2008, 2013, 2016, 2019 |
| South Africa | 2016 |
| Tanzania | 2007-2008, 2010, 2011-2012, 2015-2016, 2017 |
| Togo | 2013-2014, 2017 |
| Uganda | 2006, 2009, 2011, 2014-2015, 2016, 2018-2019 |
| Zambia | 2007, 2013-2014, 2018 |
| Zimbabwe | 2005-2006, 2010-2011, 2015 |

**Supplemental Table 2.** Common Pathogens in Africa and their associated incubation periods. Pathogen Illnesses and Deaths are taken from a study by Thystrup et al. (2024).

| Pathogen | Type | Estimated Incubation Time | Whole African Region (197 mil. Pop) |  | Zoonotic Potential<sup>%</sup> |
| --- | --- | --- | --- | --- | --- |
|  |  |  | Illnesses (95% UI) | Deaths (95% UI) |  |
| Rotavirus | Virus | 1-2 Days <sup>^</sup> | 9,457,547 (4,931,180–13,115,905) | 27,616 (18,226–45,459) | Weak |
| Norovirus | Virus | 12-48 Hours <sup>^</sup> | 10,257,653 (4,650,470–14,555,680) | 9,135 (5,980–15,005) | Weak |
| Astrovirus | Virus | 4-5 days <sup>^</sup> | 2,544,662 (1,196,629-3,590,575) | 457 (299–750) | Weak |
| Cryptosporidium spp. | Protozoan | 2-10 days (usually 7 days) <sup>§</sup> | 5,901,845 (2,695,492-8,365,420) | 22,121 (14,525–36,363) | Strong |
| Entamoeba histolytica | Protozoan | 2-4 weeks <sup>§</sup> | 883,095 (418,536-1,244,527) | 1,802 (1,161-2,948) | Weak |
| Giardia lamblia | Protozoan | 1-14 days (usually 7 days) <sup>§</sup> | 14,037,258 (6,437,064–19,884,469) | 9,198 (6,079–15,148) | Strong |
| Campylobacter spp. | Bacteria | 1-10 days (usually 2-4 days) <sup>§</sup> | 5,902,020 (2,611,903-8,405,257) | 8,108 (5,271–13,293) | Strong |
| Enterohemorrhagic E. coli (EHEC): | Bacteria | 2-5 days <sup>&</sup> | 2,919,966 (1,266,400-4,170,655) | 5,076 (3,290-8,315) | Strong |
| Enteropathogenic E. coli (EPEC): | Bacteria | 9-12 hours <sup>§</sup> | 7,517,917 (3,370,830–10,685,772) | 12,317 (8,037–20,214) | Strong |
| Enterotoxigenic E. coli (ETEC): | Bacteria | 10-72 hours<sup>§</sup> | 10,044,953 (4,476,717–14,290,494) | 6,988 (4,607–11,500) | None |
| Enteraggregative E. coli (EAEC): | Bacteria | 8-48 hours<sup>§</sup> | 17,137,586 (7,924,333–24,245,224) | 21,471 (14,013–35,237) | Weak |
| Enteroinvasive E. coli (EIEC): | Bacteria | 10-18 hours<sup>§</sup> | 3,961,764 (2,027,562-5,512,315) | 70,108 (46,172–115,341) | Weak |
| Shiga toxin-producing E. coli (STEC): | Bacteria | 1–10 days (usu. 3–4 days) <sup>§</sup> | 8,468,956 (4,086,157–11,900,853) | 633 (428-1,050) | Strong |
| V. cholerae | Bacteria | 1-2 days<sup>*</sup> | 76,033 (43,923–103,420) | 3,968 (2,697-6,586) | Weak |
| Salmonella spp. | Bacteria | 12 hours - 4 days<sup>^</sup> | 889,172 (422,620-1,252,524) | 700 (479-1,163) | Strong |
| Shigella spp. | Bacteria | 1-2 days<sup>§</sup> | 2,050,246 (971,448-2,889,486) | 10,596 (6,929–17,398) | Weak |

<sup>%</sup> Pathogens were classified into the following categories based on currently available research: strong zoonotic potential, weak or rare zoonotic potential (especially if zoonotic transmission is not the primary transmission pathway), and no zoonotic potential.
<sup>^</sup> Source: Cleveland Clinic
<sup>§</sup> Source: CDC Yellow Book
<sup>\*</sup> Azman et al., 2014
<sup>&</sup> Hopkins Medicine

**Supplemental Table 3.**
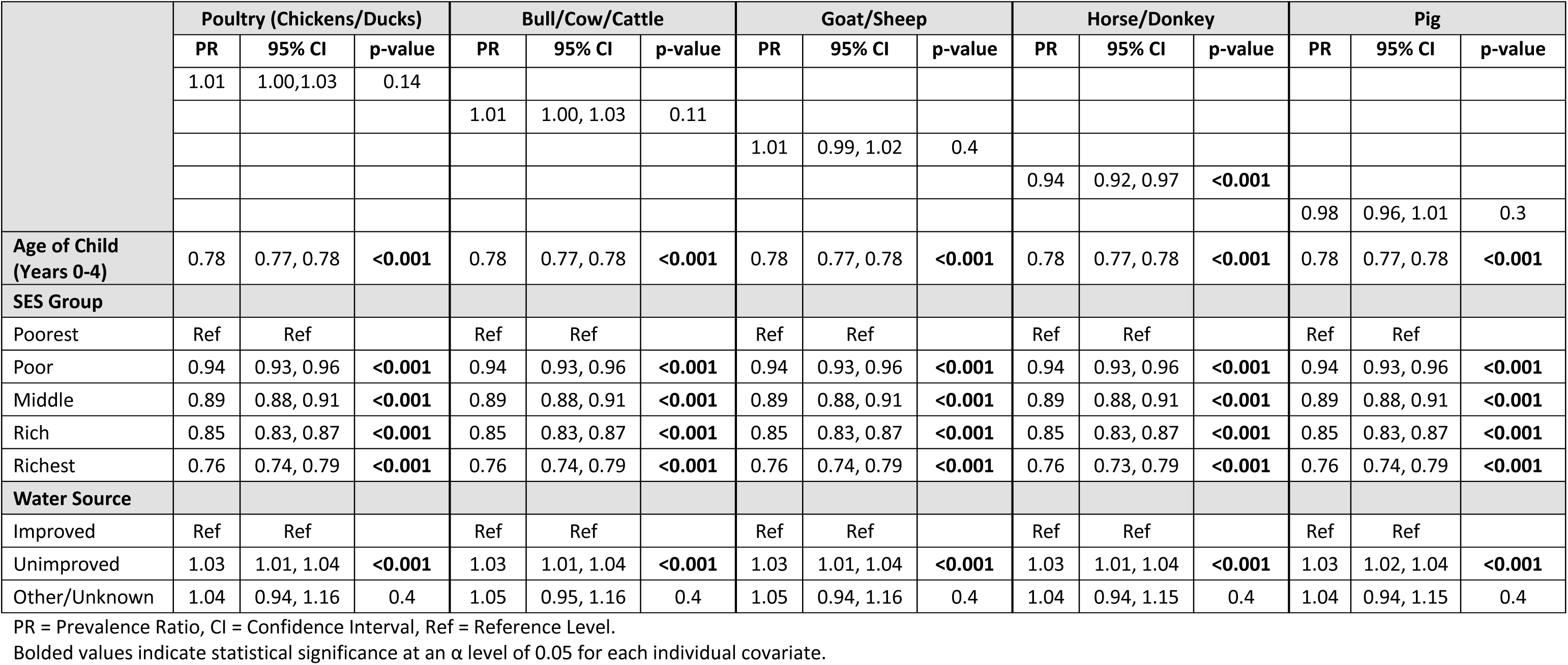
Model results from the mixed-effects Poisson regression models with a log link when stratifying by the types of animals that a family owns. The outcome of these models is a prevalence ratio of children with two-week diarrhea compared to children that did not experience two-week diarrhea.

|  | Poultry (Chickens/Ducks) |  |  | Bull/Cow/Cattle |  |  | Goat/Sheep |  |  | Horse/Donkey |  |  | Pig |  |  |
| --- | --- | --- | --- | --- | --- | --- | --- | --- | --- | --- | --- | --- | --- | --- | --- |
|  | PR | 95% CI | p-value | PR | 95% CI | p-value | PR | 95% CI | p-value | PR | 95% CI | p-value | PR | 95% CI | p-value |
|  | 1.01 | 1.00,1.03 | 0.14 |  |  |  |  |  |  |  |  |  |  |  |  |
|  |  |  |  | 1.01 | 1.00, 1.03 | 0.11 |  |  |  |  |  |  |  |  |  |
|  |  |  |  |  |  |  | 1.01 | 0.99, 1.02 | 0.4 |  |  |  |  |  |  |
|  |  |  |  |  |  |  |  |  |  | 0.94 | 0.92, 0.97 | <0.001 |  |  |  |
|  |  |  |  |  |  |  |  |  |  |  |  |  | 0.98 | 0.96, 1.01 | 0.3 |
| **Age of Child (Years 0-4)** | 0.78 | 0.77, 0.78 | <0.001 | 0.78 | 0.77, 0.78 | <0.001 | 0.78 | 0.77, 0.78 | <0.001 | 0.78 | 0.77, 0.78 | <0.001 | 0.78 | 0.77, 0.78 | <0.001 |
| **SES Group** |  |  |  |  |  |  |  |  |  |  |  |  |  |  |  |
| Poorest | Ref | Ref |  | Ref | Ref |  | Ref | Ref |  | Ref | Ref |  | Ref | Ref |  |
| Poor | 0.94 | 0.93, 0.96 | <0.001 | 0.94 | 0.93, 0.96 | <0.001 | 0.94 | 0.93, 0.96 | <0.001 | 0.94 | 0.93, 0.96 | <0.001 | 0.94 | 0.93, 0.96 | <0.001 |
| Middle | 0.89 | 0.88, 0.91 | <0.001 | 0.89 | 0.88, 0.91 | <0.001 | 0.89 | 0.88, 0.91 | <0.001 | 0.89 | 0.88, 0.91 | <0.001 | 0.89 | 0.88, 0.91 | <0.001 |
| Rich | 0.85 | 0.83, 0.87 | <0.001 | 0.85 | 0.83, 0.87 | <0.001 | 0.85 | 0.83, 0.87 | <0.001 | 0.85 | 0.83, 0.87 | <0.001 | 0.85 | 0.83, 0.87 | <0.001 |
| Richest | 0.76 | 0.74, 0.79 | <0.001 | 0.76 | 0.74, 0.79 | <0.001 | 0.76 | 0.74, 0.79 | <0.001 | 0.76 | 0.73, 0.79 | <0.001 | 0.76 | 0.74, 0.79 | <0.001 |
| **Water Source** |  |  |  |  |  |  |  |  |  |  |  |  |  |  |  |
| Improved | Ref | Ref |  | Ref | Ref |  | Ref | Ref |  | Ref | Ref |  | Ref | Ref |  |
| Unimproved | 1.03 | 1.01, 1.04 | <0.001 | 1.03 | 1.01, 1.04 | <0.001 | 1.03 | 1.01, 1.04 | <0.001 | 1.03 | 1.01, 1.04 | <0.001 | 1.03 | 1.02, 1.04 | <0.001 |
| Other/Unknown | 1.04 | 0.94, 1.16 | 0.4 | 1.05 | 0.95, 1.16 | 0.4 | 1.05 | 0.94, 1.16 | 0.4 | 1.04 | 0.94, 1.15 | 0.4 | 1.04 | 0.94, 1.15 | 0.4 |

PR = Prevalence Ratio, CI = Confidence Interval, Ref = Reference Level.
Bolded values indicate statistical significance at an $\alpha$ level of 0.05 for each individual covariate.

**Supplemental Table 4.** Model results from the mixed-effects Poisson regression models with a log link for every 5, 10 and 15 animals that a family owns. The outcome of these models is a prevalence ratio of children with two-week diarrhea compared to children that did not experience two-week diarrhea.

|  | Every 5 Animals Owned |  |  | Every 10 Animals Owned |  |  | Every 15 Animals Owned |  |  |
| --- | --- | --- | --- | --- | --- | --- | --- | --- | --- |
|  | PR | 95% CI | p-value | PR | 95% CI | p-value | PR | 95% CI | p-value |
|  | 1.00 | 0.99, 1.00 | <0.001 |  |  |  |  |  |  |
|  |  |  |  | 0.99 | 0.99, 1.00 | <0.001 |  |  |  |
|  |  |  |  |  |  |  | 0.99 | 0.98, 0.99 | <0.001 |
| **Age of Child (Years 0-4)** | 0.78 | 0.77, 0.78 | <0.001 | 0.78 | 0.77, 0.78 | <0.001 | 0.78 | 0.77, 0.78 | <0.001 |
| **SES Group** |  |  |  |  |  |  |  |  |  |
| Poorest | Ref | Ref |  | Ref | Ref |  | Ref | Ref |  |
| Poor | 0.94 | 0.93, 0.96 | <0.001 | 0.94 | 0.93, 0.96 | <0.001 | 0.94 | 0.93, 0.96 | <0.001 |
| Middle | 0.89 | 0.88, 0.91 | <0.001 | 0.89 | 0.88, 0.91 | <0.001 | 0.89 | 0.88, 0.91 | <0.001 |
| Rich | 0.85 | 0.83, 0.87 | <0.001 | 0.85 | 0.83, 0.87 | <0.001 | 0.85 | 0.83, 0.87 | <0.001 |
| Richest | 0.76 | 0.74, 0.79 | <0.001 | 0.76 | 0.74, 0.79 | <0.001 | 0.76 | 0.74, 0.79 | <0.001 |
| **Water Source** |  |  |  |  |  |  |  |  |  |
| Improved | Ref | Ref |  | Ref | Ref |  | Ref | Ref |  |
| Other/Unknown | 1.04 | 0.94, 1.15 | 0.4 | 1.04 | 0.94, 1.15 | 0.4 | 1.04 | 0.94, 1.16 | 0.4 |
| Unimproved | 1.03 | 1.02, 1.04 | <0.001 | 1.03 | 1.02, 1.04 | <0.001 | 1.03 | 1.02, 1.04 | <0.001 |

